# Gut bacterial tyrosine decarboxylase gene abundance associates with disease duration, medication exposure, and gastrointestinal symptoms in a longitudinal cohort of Parkinson’s disease patients

**DOI:** 10.1101/2021.07.13.21259300

**Authors:** Sebastiaan P. van Kessel, Petri Auvinen, Filip Scheperjans, Sahar El Aidy

**Author notes:** Correspondence Filip Scheperjans and Sahar El Aidy. shared last author (Filip Scheperjans, Sahar El Aidy).

## Abstract

Gut microbiota influences the clinical response of a wide variety of orally administered drugs. However, the underlying mechanisms by which drug-microbiota interactions occur are still obscure. Previously, we reported that tyrosine decarboxylating (TDC) bacteria may restrict the levels of levodopa reaching the circulation in patients with Parkinson’s disease (PD). We observed a significant positive association between disease duration and the abundance of the bacterial *tdc*-gene. The question arises whether increased exposure to anti-PD medication could affect the abundance of bacterial TDC, to ultimately impact drug efficacy. To this end, we investigated the potential association between anti-PD drug exposure and bacterial *tdc*-gene abundance over a time period of two years in a longitudinal cohort of PD patients and healthy controls. Our data reveal significant associations between *tdc*-gene abundance, anti-PD medication, and gastrointestinal symptoms and warrants further research on the effect of anti-PD medication on microbial changes and gastrointestinal-function.

## Introduction

In recent years, many studies focused on the changes in the microbiota composition in individuals with Parkinson’s disease (PD) compared to healthy subjects (extensively covered in several (systematic) reviews (Boertien et al., 2019; van Kessel and El Aidy, 2019a among others). While certain differential abundance alterations were reproduced across multiple studies, variation of results across studies was considerable (Boertien et al., 2019; van Kessel and El Aidy, 2019a).

One of the reasons that may explain the inconsistency among these studies, are confounding factors such as anti-PD medications, disease duration, and GI symptoms. Indeed, studies took these factors into account with variable effort. Catechol-O-methyltransferase (COMT) inhibitors, anticholinergic and potentially levodopa/carbidopa were found to have a significant effect on the changes in the microbiota profile (Aho et al., 2019; Hill-Burns et al., 2017; Scheperjans et al., 2015; Weis et al., 2019).

Besides medication, GI-dysfunction should be considered when analyzing the altered microbiota in PD patients. Indeed, PD patients usually experience more GI-dysfunction symptoms compared to healthy controls (HC) (Fasano et al., 2015; Kenna et al., 2021) and intestinal transit time can impact microbiota composition (Falony et al., 2016).

Moreover, it has been shown that there is an association between anti-PD medications and GI symptoms. For example, anti-PD medications were shown to have associations (corrected for disease duration) with the total GI Symptoms Rating Score, upper GI symptoms and hypoactive GI functions (Kenna et al., 2021). Furthermore, the proportion of the COMT inhibitor usage was significantly higher in patients with an abnormal transit compared to those with normal transit (Khoshbin et al., 2020). However, the statistical analysis in that study could not distinguish whether the levodopa equivalent daily dose (LEDD) or disease duration was the more contributing factor to the slow colon transit (Khoshbin et al., 2020). In addition, *ex vivo* rodent studies and *in vivo* dog and human studies showed an effect of dopamine agonists and/or dopamine (which can originate from levodopa in PD patients) on the gut motility, recently reviewed (van Kessel and El Aidy, 2019b, and citations in there). Gut microbial metabolization of unabsorbed residues of levodopa were also shown to influence ileal-motility *ex vivo* (van Kessel et al., 2020).

Recent studies have shown that tyrosine decarboxylating (TDC) bacteria can decarboxylate levodopa into dopamine in the periphery, thereby may restrict the levels of levodopa available for the brain (van Kessel et al., 2019; Maini Rekdal et al., 2019). Potentially, TDC-harboring bacteria could create a vicious-circle, where peripheral dopamine production affects the gut motility, favoring the colonization of (TDC)-bacteria (van Kessel et al., 2019). Additionally, non-levodopa anti-PD medications (Monoaminoxidase inhibitors, COMT inhibitors, and dopamine agonists), which affect the peripheral dopaminergic-balance, may lead to an altered GI-function, potentially leading to an overgrowth of (TDC)-bacteria, ultimately affecting the bioavailability of levodopa. However, levels of TDC-bacteria have not been measured nor were previously correlated with GI symptoms in longitudinal PD cohorts.

In this study we focused on measuring fecal *tdc*-gene abundance and its association with anti-PD medication exposure in a 2 year longitudinal cohort consisting of 67 PD and 65 healthy matched subjects, that was used previously for investigating microbiota and PD (Aho et al., 2019; Scheperjans et al., 2015).

## Material and Methods

### Cohort

The original gender, age and sex matched cohort was recruited for a pilot study in 2015 investigating PD and gut microbiota (Scheperjans et al., 2015). All subjects were invited to a follow-up on average 2.25±0.20 years later to investigate temporal stability in the PD microbiota (Aho et al., 2019). The study was approved by the ethics committee of the Hospital District of Helsinki and Uusimaa. All participants gave written informed consent and the study was registered at clinicaltrials.gov (NCT01536769).

From the total 165 subjects (77 PD, 88 HC) recruited in the baseline and follow-up studies (Aho et al., 2019; Scheperjans et al., 2015), 13 subjects (6 PD, 7 HC) were excluded because they did not return for the follow-up study, and 20 subjects (4 PD, 16 HC) were excluded because of various other reasons at baseline or follow up. In the control group, 1 subject was excluded for a sibling with PD, 3 subjects for having a common cold, 8 subjects for hyposmia (pre-motor PD symptom), 2 subjects had a recent surgery, 1 subject had no matching sample, and 1 sample was missing. In the PD group, 1 subject was excluded because of recent surgery, 1 subject had a change in diagnosis, 1 subject because of a sampling handling issue, 1 subject because of medical history. In total 33 subjects (10 PD, 23 HC) were excluded, resulting in 132 subjects (67 PD, 65 HC) used in this study.

The following parameters were assessed as described in the previous studies (Scheperjans, 2015, Aho, 2019): Gastrointestinal symptoms (Wexner constipation score (Agachan et al., 1996), Rome III questionnaire (Longstreth et al., 2006)), disease severity (Unified Parkinson’s Disease Rating Scale (UPDRS) (Fahn, S., Elton, R., and Members of the UPDRS Development Committee, 1987), and medication exposure.

### DNA extraction

Stool sample collection and DNA isolation was performed in a previous study (Aho et al., 2019). Briefly, stool samples were collected by the study subjects into collection tubes with pre-filled DNA stabilizer (PSP Spin Stool DNA Plus Kit, STRATEC Molecular), and stored in the refrigerator until transport (for up to 3 days). After receival of the samples, they were transferred to −80 °C. DNA from both baseline and follow-up samples were extracted with the PSP Spin Stool DNA Plus Kit (STRATEC Molecular). Each extraction batch included one blank sample to assess potential contamination. (Of note, to prevent potential technical differences, DNA form the baseline samples were extracted at the baseline (Scheperjans et al., 2015) and at follow-up study (Aho et al., 2019), thus the baseline samples were thawed twice.)

### Determination of *tdc*-gene abundance

The DNA concentration of the samples was directly estimated from the 96-well plates by measuring the (pathlength corrected) absorbance at 260 nm and 320 nm in a multimode reader, the DNA concentration was calculated as followed: 50 × (sample^260-320^ − blank^260-320^). Samples that were negative, very low, or very high in concentration were measured with the nanodrop to confirm. All DNA samples were diluted 20× so that the concentration range would fall in a range of 2-25 ng/µl (median, 13.7 ng/µL, interquartile range, 6.9 - 21.8 ng/µl) and 2 µl was used for quantitative PCR (qPCR). qPCR of *tdc* genes was performed using primers Dec5f (5’-CGTTGTTGGTGTTGTTGGCACNACNGARGARG-3’) and Dec3r (5’-CCGCCAGCAGAATATGGAAYRTANCCCAT-3’) targeting a 350 bp region of the *tdc* gene (Torriani et al., 2008). Primers targeting 16S rRNA gene for all bacteria (Fierer et al., 2005), Eub338 (5’-ACTCCTACGGGAGGCAGCAG-3’) and Eub518 (5’-ATTACCGCGGCTGCTGG-3’) were used as an internal control. All qPCR experiments were performed in a Bio-Rad CFX96 RT-PCR system (Bio-Rad Laboratories, Veenendaal, The Netherlands) with iQ SYBR Green Supermix (170-8882, Bio-Rad) in 10 μL reactions using the manufacturer’s protocol. qPCR was performed using the following parameters: 3 min at 95 °C; 15 sec at 95 °C, 1 min at 58 °C, 40 cycles. A melting curve was determined at the end of each run to verify the specificity of the PCR amplicons. Data analysis was performed using Bio-Rad CFX Manager 3.1 software. Ct[DEC] values were corrected with the internal control (Ct[16s]) and linearized using 2^-(Ct^[DEC]^-Ct^[16s]^) based on the 2^^-ΔΔCt^ method (Livak and Schmittgen, 2001).

### Statistics

All statistical tests were performed in IBM SPSS Statistics version 26. The p-value adjustments were performed in R version 4.0.0 using p.adjust (p-values, “fdr”). The qPCR data were tested for outliers per group and timepoint using the ROUT method (Q=0.1%) in GraphPad Prism v7 and the identified outliers were removed. All variables were tested for normality using Kolmogorov-Smirnov and Shapiro-Wilk tests using the Explore function in SPSS. Based on the distribution of the data, the differences were tested using the appropriate statistical tests. The general linear models (GLMs) were performed using the Generalized Linear Models function in SPSS and the main effects were tested using the Wald Chi Square test. Additionally, the variance inflation factor (VIF) was computed to check for potential collinearity between variables.

## Results

### Clinical variables

Comparing the clinical variables between the longitudinal cohort of PD and HC did not reveal any significant differences in sex, age (at stool collection) and BMI, no systemic antibiotics were used within the last month (**Supplementary Table 1**). The duration of motor and non-motor symptom onset in the PD cohort at baseline was ∼8 years (**Supplementary Table 1**). Over time (between baseline and follow-up), the levodopa equivalent daily dose (LEDD) significantly increased by an average of 116 mg (**Table 1**). On average, the UPDRS I and II scores significantly increased while UPDRS III (at ON-state) significantly decreased over time, respectively (**Supplementary Table 2**). The latter may be explained by the significant LEDD increase over time. The Hoehn & Yahr (at ON-state) score slightly increased over time (**Supplementary Table 2**).

**Table 1.**
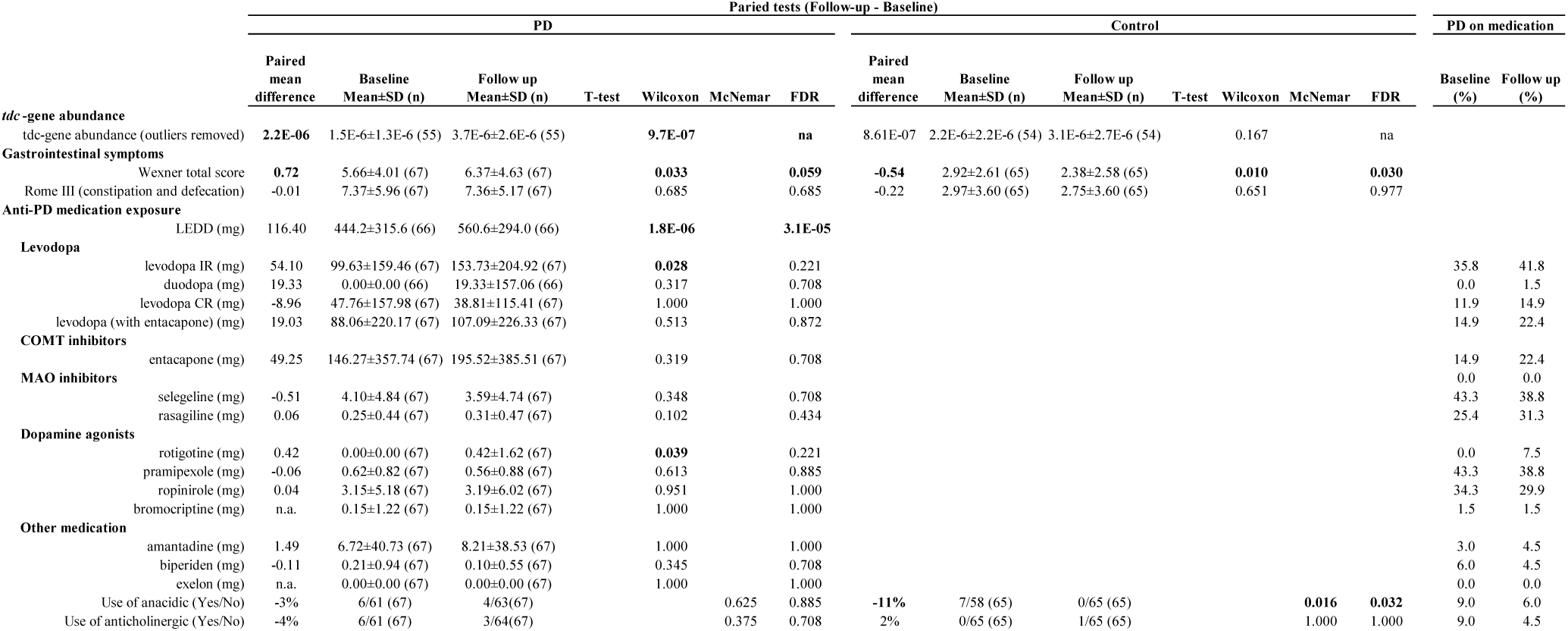
Paired tests between follow-up and baseline samples in PD and HC of tdc-gene abundance, gastrointestinal symptoms and medication. Significant test-results are printed in bold.

### The gut bacterial *tdc*-gene abundance, GI-symptoms, and medication exposure significantly increased over time in PD patients

Recently, it has be shown that TDC-bacteria in the GI-tract interfere with the availability of levodopa medication in animal models and that longer disease duration and exposure to levodopa may further increase the abundance of TDC-bacteria in the gut (van Kessel et al., 2019). Thus, we sought to investigate the changes in the levels of gut bacterial *tdc*-gene abundance over time in the longitudinal PD cohort including the differences between PD patients and matching HC.

When comparing PD patients and HC, PD patients tended to have a higher *tdc*-gene abundance (p=0.057) at follow-up (**Figure 1 and Table 2**). Correspondingly, the increase in *tdc*-gene abundance over time, was significantly higher in PD patients compared to HC subjects (Wilcoxon-test, p=9.7E-07) with a mean increase of 2.6-fold (**Table 1 and Figure 1**). The results indicate that, over time, *tdc*-gene abundance increases more rapidly in PD patients compared to HC subjects.

**Figure 1.**
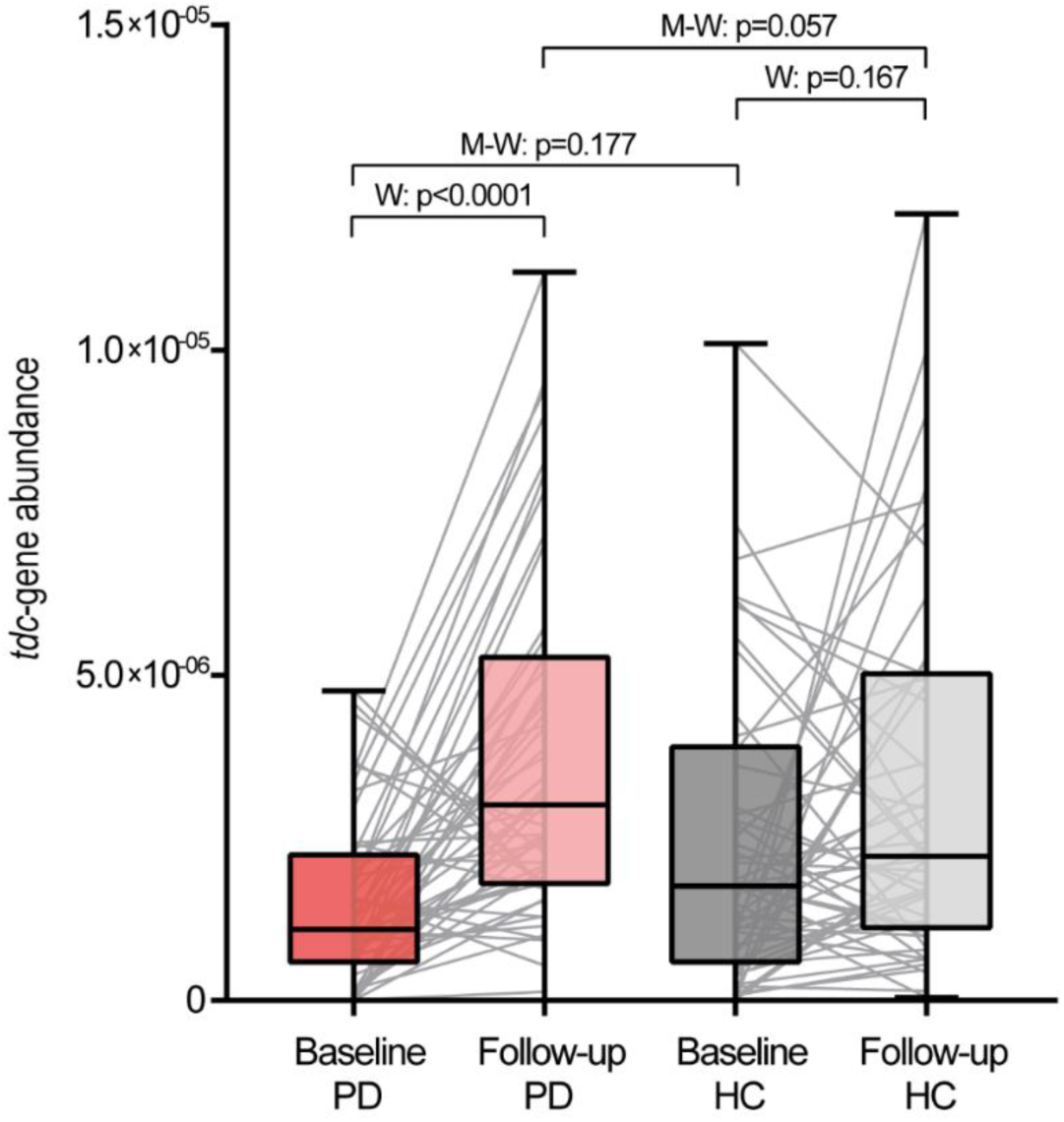
*tdc*-gene abundance in PD and healthy control subjects. The *tdc*-gene abundance is depicted for PD patients (PD, red boxes; dark, baseline; light, follow-up) and healthy control subjects (HC, grey boxes; dark, baseline; light, follow-up) for both time points. Nonparametric paired Wilcoxon tests (W) were performed to test for significant increase over time between paired samples (grey lines). Significant outliers were removed using the ROUT method (Q=0.1%). Nonparametric unpaired Mann-Whitney tests (M-W) were performed to test for significant differences between PD and HC at baseline and follow-up. Boxes represent the median with the interquartile range and whiskers the maxima and minima.

**Table 2.** Independent tests between PD and HC of *tdc*-gene abundance, gastrointestinal symptoms and medication. Significant test-results are printed in bold.

|  | Independent-tests (PD - Control) |  |  |  |  |  |  |  |  |  |  |  |  |  |
| --- | --- | --- | --- | --- | --- | --- | --- | --- | --- | --- | --- | --- | --- | --- |
|  | Baseline |  |  |  |  |  | Follow up |  |  |  |  |  |  |  |
|  | Mean Difference | PD Mean±SD (n) | Control Mean±SD (n) | T-test | Mann-Whitney | Fisher's test | FDR | Mean Difference | PD Mean±SD (n) | Control Mean±SD (n) | T-test | Mann-Whitney | Fisher's test | FDR |
| *tdc*-gene abundance |  |  |  |  |  |  |  |  |  |  |  |  |  |  |
| tdc-gene abundance (outliers removed) | -7.4E-07 | 1.5E-6±1.3E-6 (55) | 2.2E-6±2.2E-6 (54) |  | 0.177 |  | na | 6.47E-07 | 3.8E-6±2.6E-6 (57) | 3.1E-6±2.7E-6 (55) |  | 0.057 |  | na |
| **Gastrointestinal symptoms** |  |  |  |  |  |  |  |  |  |  |  |  |  |  |
| Wexner total score | **2.73** | 5.66±4.01 (67) | 2.92±2.61 (65) |  | **2.4E-05** |  | **2.4E-05** | **3.99** | 6.37±4.63 (67) | 2.38±2.58 (65) |  | **5.9E-09** |  | **1.2E-08** |
| Rome III (constipation and defecation) | **4.40** | 7.37±5.96 (67) | 2.97±3.60 (65) |  | **1.2E-06** |  | **2.4E-06** | **4.60** | 7.36±5.17 (67) | 2.75±3.60 (65) |  | **1.3E-08** |  | **1.3E-08** |
| **Medication** |  |  |  |  |  |  |  |  |  |  |  |  |  |  |
| Use of anacidic (Yes/No) | -2% | 6/61 (67) | 7/58(65) |  |  | 0.777 | 0.777 | 6% | 4/63 (67) | 0/65 (65) |  |  | 0.119 | 0.238 |
| Use of anticholinergic (Yes/No) | **9%** | 6/61 (67) | 0/65 (65) |  |  | **0.028** | **0.056** | 3% | 3/64 (67) | 1/64 (65) |  |  | 0.619 | 0.619 |

Because the GI transit time also impacts the microbial composition (Falony et al., 2016), including the TDC bacteria, differences in GI-symptoms were assessed at baseline and follow-up. At both time points, the GI symptoms were significantly more severe in PD patients compared to HC subjects (**Table 2**).

Only the Wexner scores, but not the Rome III scores, increased significantly over time in PD patients while in HC subjects, the Wexner scores decreased significantly over time (**Table 1**).

Although the LEDD increased significantly over time (**Table 1**), no significant increase was observed for any individual drug in PD patients after correction for FDR, possibly due to the changes of the type of medication (**Table 1**). Nonetheless, at baseline, anticholinergic medication use was significantly higher in PD patients compared to HC subjects (**Table 2**). Only in the HC group, a significant decrease in anacidic medication use was observed over time (**Table 1**).

### Anti-PD medications and GI symptoms associate with the gut bacterial *tdc*-gene abundance over time

Using general linear models (GLM), the contribution of the difference in anti-PD medication exposure to the difference in *tdc*-gene abundance over time (follow-up − baseline) was assessed (**Table 3**). The model showed that dose changes of entacapone, rasagiline, pramipexole, and ropinirole significantly contributed to the difference in *tdc*-gene abundance over time. Entacapone and the dopamine agonists contributed positively to the difference in *tdc*-gene abundance, while MAOi contributed negatively to the *tdc*-gene abundance over time, respectively.

**Table 3.**
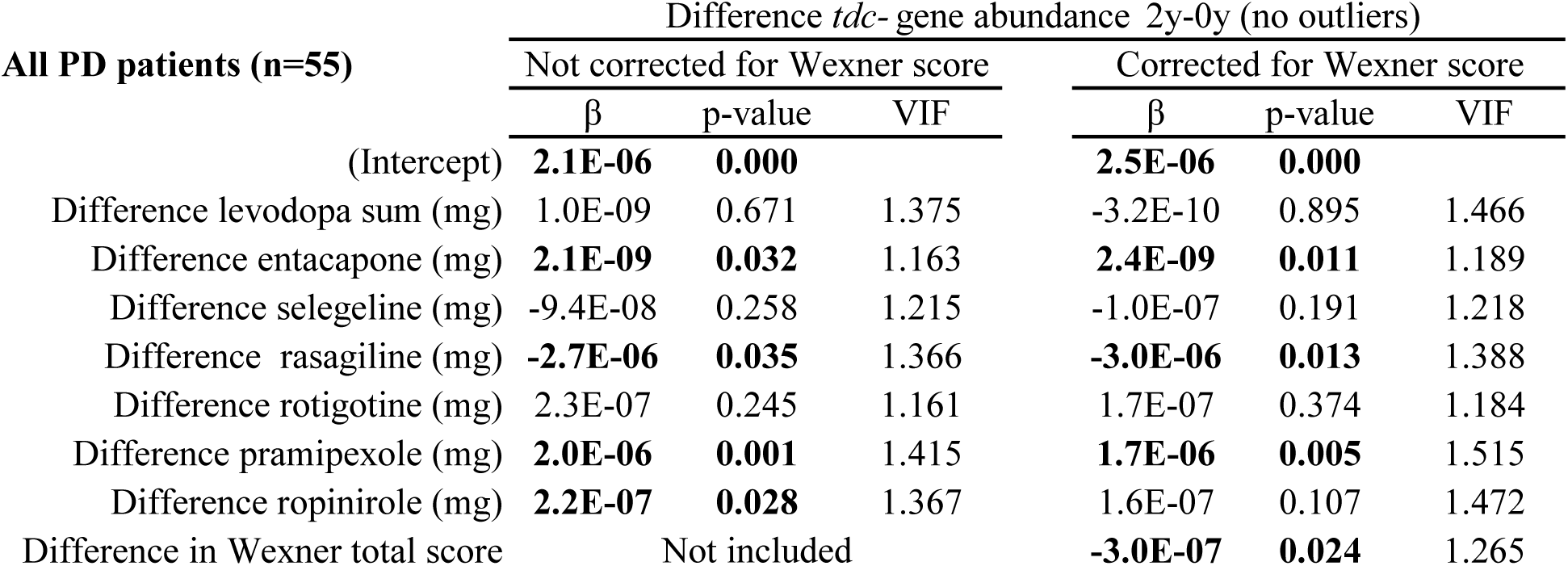
General linear model of the difference *tdc*-gene abundance overtime with anti-PD medication and Wexner-score as variables. Significant variable contributions to the model are printed in bold. VIF; Variance Inflation Factor.

| All PD patients (n=55) | Difference *tdc*- gene abundance 2y-0y (no outliers) |  |  |  |  |  |
| --- | --- | --- | --- | --- | --- | --- |
|  | Not corrected for Wexner score |  |  | Corrected for Wexner score |  |  |
| | $\beta$ | p-value | VIF | $\beta$ | p-value | VIF |
| (Intercept) | **2.1E-06** | **0.000** |  | **2.5E-06** | **0.000** |  |
| Difference levodopa sum (mg) | 1.0E-09 | 0.671 | 1.375 | -3.2E-10 | 0.895 | 1.466 |
| Difference entacapone (mg) | **2.1E-09** | **0.032** | 1.163 | **2.4E-09** | **0.011** | 1.189 |
| Difference selegeline (mg) | -9.4E-08 | 0.258 | 1.215 | -1.0E-07 | 0.191 | 1.218 |
| Difference rasagiline (mg) | **-2.7E-06** | **0.035** | 1.366 | **-3.0E-06** | **0.013** | 1.388 |
| Difference rotigotine (mg) | 2.3E-07 | 0.245 | 1.161 | 1.7E-07 | 0.374 | 1.184 |
| Difference pramipexole (mg) | **2.0E-06** | **0.001** | 1.415 | **1.7E-06** | **0.005** | 1.515 |
| Difference ropinirole (mg) | **2.2E-07** | **0.028** | 1.367 | 1.6E-07 | 0.107 | 1.472 |
| Difference in Wexner total score |  | Not included |  | **-3.0E-07** | **0.024** | 1.265 |

Because the Wexner scores, but not Rome III, significantly increased over time in the PD group (**Table 1**), this factor was included in the model to correct for its potential contribution to the *tdc*-gene abundance. Remarkably, Wexner total scores significantly contributed negatively to the *tdc*-gene abundance (**Table 3**), suggesting that subjects with less constipation have an increased *tdc*-gene abundance. Correction for Wexner scores showed that the difference in exposure to anti-PD medication, stated above, still contributed to the model except for ropinirole (p=0.107). The results indicate that prolonged exposure of these specific anti-PD medications, excluding levodopa, contributed to *tdc*-gene abundance independent of the changes in GI symptoms measured by Wexner scores.

PD patients usually require change in the anti-PD dosage regimen during the disease progression, compared to patients in a steady state of the disease. Thus, we sought to investigate whether the differences in anti-PD dosage regimen between the two groups was the factor contributing to the changes in *tdc-*gene abundance observed above (**Table 3**). To this end, the PD-group was sub-divided into steady (n=35) and progressing (n=12) PD patients as described and performed in Aho *et al*. (Aho et al., 2019). Comparing the mean differences of the medications of the stable and progressing PD patients over time showed that exposure to levodopa and entacapone significantly increased, while pramipexole exposure significantly decreased in the progressing group compared to the steady group (**Table 4**).

**Table 4.**
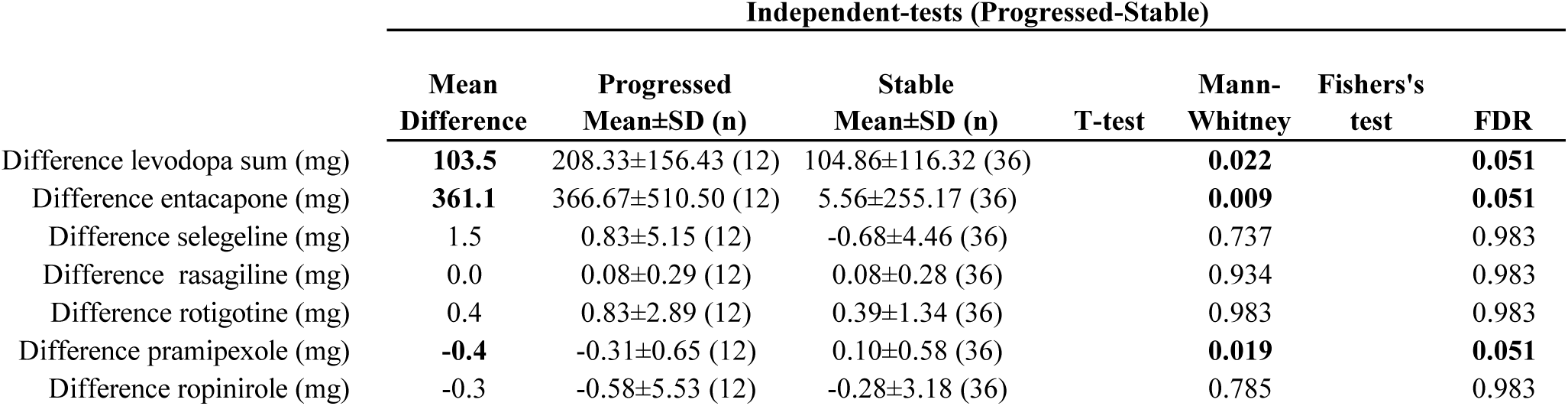
Independent tests between progressed and stable PD patients of exposure of anti-PD medications overtime. Significant test-results are printed in bold.

When comparing the steady PD patients group with the progressed PD patients group (**Table 5**) only entacapone was not associated with *tdc*-gene abundance and rotigotine now significantly contributed to the model (which was not observed in all the PD patients, **Table 3**), however the significance was lost when correcting for Wexner-score.

**Table 5.**
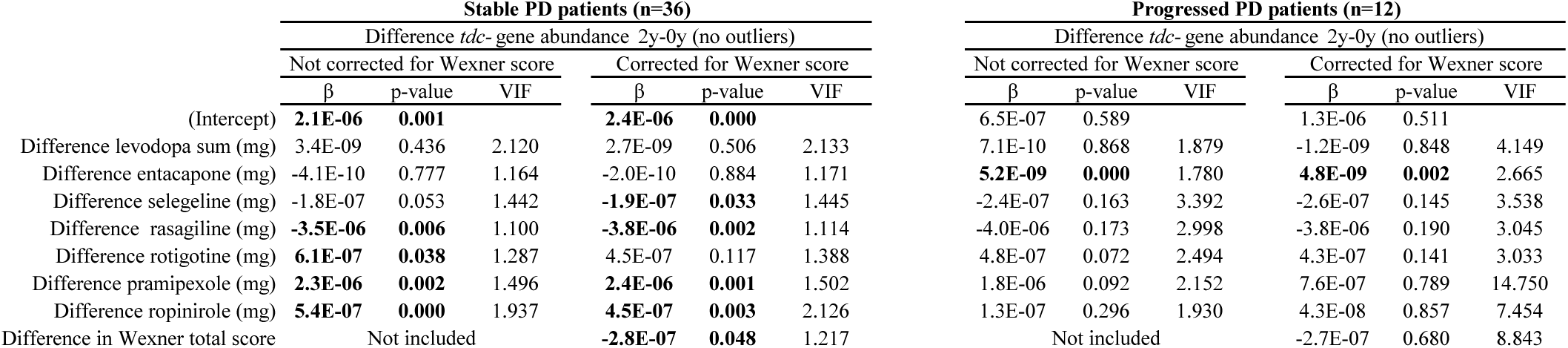
General linear model of the difference of *tdc*-gene abundance overtime with anti-PD medication and Wexner-score as variable in steady and progressing PD patients.□Significant variable contributions to the model are printed in bold. VIF; Variance Inflation Factor.

In the progressing PD group, only entacapone contributed significantly to the change in *tdc*-abundance (**Table 5**). Because the variation inflation factor (VIF, which tests if the variance of a variable increases with another) suggested collinearity between the factors in the progressing PD group, DA-agonists and MAO-inhibitors were combined using LED calculation (Tomlinson et al., 2010). Using the combined variables in the GLM, no collinearity was observed anymore, while entacapone still contributed significantly to the *tdc*-gene abundance (**Supplementary Table 3**). These results indicate that the difference in drug exposure over time between stable and progressed PD patients (**Table 4**), reflect their contribution to the *tdc*-gene abundance in the GLMs (**Table 5 and Supplementary Table 3**). In summary these observations show that, likely due to the change in exposure to these specific anti-PD medications, entacapone is the significant factor contributing to the increase in the *tdc-*gene abundance in progressing PD patients, while the other anti-PD medications contribute to the *tdc*-gene abundance in stable PD patients.

## Discussion and Conclusion

In this study, we have established that gut bacterial *tdc*-gene abundance is significantly increasing over time in PD patients (**Table 1**), in line with previous results, where a significant correlation between the disease duration and the *tdc*-gene abundance was observed (van Kessel et al., 2019). The levels of gut bacterial *tdc*-gene abundance were not significantly different compared to HC at baseline, but close to significant at follow-up (**Table 2**). Accordingly, the increase in *tdc*-gene abundance was 2.6-fold higher in PD than HC, suggesting that that the increased gene abundance occurs more rapidly in PD patients. In this study we did not find a significant correlation between the levodopa dosages and the *tdc*-gene abundance. This discrepancy could be explained by the relative lower proportion of high levodopa dosages in this study. At baseline and follow-up 19.4% (max 900 mg) and 26.9% (max 875 mg) of the PD patients had a dose higher than 400 mg/day respectively, while in the previous study (van Kessel et al., 2019) 60% of the PD patients received a dosage higher than 400 mg/day (max 1100 mg).

Using GLMs, we showed that other anti-PD medications than levodopa contributed significantly to the *tdc*-gene abundance. Importantly, all the tested medications (**Table 3**) affect the (peripheral) dopaminergic-system; COMT inhibitors prevent methylation of levodopa, dopamine, and norepinephrine; MAO inhibitors prevent dopamine, and norepinephrine oxidation; and DA-agonists act on dopamine receptors expressed in the gut. Collectively, these medications were recently shown to elicit an effect on the GI symptoms (Kenna et al., 2021). Although GI-dysfunction might be caused by the degeneration of enteric neurons, as observed in PD patients with chronic constipation (Singaram et al., 1995) and reported in a MPTP mouse model for PD (Anderson et al., 2007), additional dopaminergic medication may impact the GI-function even further. Indeed, the Wexner-score, which significantly increased over time in PD patients, did not change the associations between anti-PD medication and *tdc*-gene abundance (except for ropinirole exposure) when considered as a confounder. This potential link between changes in GI symptoms, as measured by Wexner score, and anti PD medications is in agreement with the outcome of a comprehensive meta-analysis showing that PD patients on ropinirole did not have higher risk of constipation compared to placebo, while those on pramipexole had a higher risk of constipation (Kulisevsky and Pagonabarraga, 2010). Unlike the Wexner score, The Rome III (constipation and defecation) score did not change over time in PD patients which may be explained by the fact that Rome III assesses symptoms retrospectively over a 3-month period which may reduce sensitivity to change. The difference observed between the two questionnaires, conforms the need of the development for more sophisticated protocols to detect and investigate GI symptoms in PD patients (Kenna et al., 2021).

Notably, only entacapone exposure in progressing PD patients contributed to the fecal *tdc*-abundance. In fact, *Enterococcus* (genus consisting of species harboring TDCs), among others were found to be significantly increased only in PD patients treated with entacapone (Weis et al., 2019). However, in their study, Weis et al. did not report whether the tested PD patients were on medications, such as MAOi or DA-agonists, other than levodopa and/or entacapone (Weis et al., 2019). Here we show that, next to entacapone, other anti-PD medications may affect gut bacterial *tdc*-gene abundance as well (**Table 3**).

The major limitation of this study is that we determined the bacterial *tdc*-gene abundance in fecal samples, which might not reflect the *tdc*-gene levels in the small-intestine, the main absorption site of levodopa and other medication. Besides, the presence of these genes do not necessarily reflect the TDC activity.

In summary, the present study implies important associations between anti-PD medication and gut bacterial *tdc*-gene abundance. These associations point towards a complex interactions between anti-PD medication, GI symptoms and gut bacterial *tdc*-gene abundance, which warrants for further research.

## Data Availability

Clinical data are not publicly available due to participant privacy and are available from the
corresponding authors on reasonable request.

## Table legends (For the actual tables see attached XLSX-file)

Table 6. General linear model of the difference of *tdc*-gene abundance over time with anti-PD medication and Wexner-score as variable in progressing PD patients (with collinearity observed). Significant variable contributions to the model are printed in bold. VIF; Variance Inflation Factor

**Supplementary Table 1.** Independent test of the basic variables Sex, Age, and BMI. Significant test-results are printed in bold.

| **Independent-tests (PD - Control)** |  |  |  |  |  |  |  |
| --- | --- | --- | --- | --- | --- | --- | --- |
|  | **Mean**<br>**Difference** | **PD**<br>**Mean±SD (n)** | **Control**<br>**Mean±SD (n)** | **T-test** | **Mann-**<br>**Whitney** | **Fishers's**<br>**test** | **FDR** |
| Sex (Female/Male) | 0.02% | 33/34 (67) | 32/33 (65) |  |  | 1.000 |  |
| BMI at baseline (kg/m<sup>2</sup>) | 0.95 | 27.2±4.3 (62) | 26.2±3.5 (61) | 0.181 |  |  |  |
| BMI at follow up (kg/m<sup>2</sup>) | 0.71 | 27.3±4.6 (67) | 26.6±3.7 (65) | 0.330 |  |  |  |
| Age at stool collection baseline (years) | 0.76 | 65.4±5.5 (67) | 64.6±7.0 (65) | 0.486 |  |  |  |
| Age at stool collection follow up (years) | 0.92 | 67.6±5.5 (66) | 66.7±6.9 (64) | 0.399 |  |  |  |
| **Age symptom onset** |  |  |  |  |  |  |  |
| Age at motor symptoms onset (years) |  | 59.5±5.3 (66) |  |  |  |  |  |
| Age at non-motor symptoms onset (years) |  | 57.7±7.8 (53) |  |  |  |  |  |
| Duration of motor symptoms (years) |  | 8.2±4.1 (66) |  |  |  |  |  |
| Duration of non-motor symptoms (years) |  | 10.0±6.9 (52) |  |  |  |  |  |

**Supplementary Table 2.** Paired tests between follow-up and baseline of LEDD, and UPDRS scores. Significant test-results are printed in bold.

| Paired-tests (Follow-up - Baseline) |  |  |  |  |  |  |  |
| --- | --- | --- | --- | --- | --- | --- | --- |
|  | Mean<br>Difference | Baseline Mean±SD<br>(n) | Follow up<br>Mean±SD (n) | T-test | Wilcoxon | McNemar | FDR |
| UPDRS I | 0.63 | 1.8±1.8 (67) | 2.5±1.8 (67) |  | **0.002** |  | **0.003** |
| UPDRS II | 1.78 | 11.6±5.5 (67) | 11.6±5.5 (67) |  | **0.001** |  | **0.003** |
| UPDRS III (ON-state) | -3.11 | 31.9±8.9 (64) | 28.8±8.7 (64) | **0.001** |  |  | **0.003** |
| UPDRS III (OFF-state) |  | na | 33.7±10.6 (65) |  |  |  |  |
| UPDRS IV | 0.84 | 2.2±2.2 (67) | 3.1±3.2 (67) |  | **0.008** |  | **0.010** |
| H&Y (ON-state) | 0.13 | 2.4±.5 (64) | 2.5±0.7 (64) |  | 0.054 |  | 0.054 |
| H&Y (OFF-state) |  | na | 2.5±0.8 (64) |  |  |  |  |

**Supplementary Table 3.** General linear model of the difference *tdc*-gene abundance over time with anti-PD medication LEDD combined and Wexner-score as variables in progressing PD patients.□.

| Progressed PD patients (n=12) | Difference *tdc*- gene abundance 2y-0y (no outliers) |  |  |  |  |  |
| --- | --- | --- | --- | --- | --- | --- |
|  | Not corrected for Wexner score |  |  | Corrected for Wexner score |  |  |
| | $\beta$ | p-value | VIF | $\beta$ | p-value | VIF |
| (Intercept) | 6.8E-07 | 0.550 |  | 1.5E-06 | 0.273 |  |
| Difference levodopa sum (mg) | -4.8E-10 | 0.913 | 1.500 | -2.3E-09 | 0.619 | 1.783 |
| Difference entacapone (mg) | **4.5E-09** | **0.001** | 1.414 | **4.6E-09** | **0.000** | 1.421 |
| Difference MAOi LED (mg) | -1.0E-08 | 0.534 | 1.315 | -9.6E-09 | 0.547 | 1.317 |
| Difference in DA agonist LED (mg) | 7.1E-09 | 0.219 | 2.383 | 3.2E-09 | 0.641 | 3.601 |
| Difference in Wexner total score |  | Not included |  | -3.4E-07 | 0.323 | 1.874 |

## Author contributions

SVK performed the qPCR, the statistical analysis, and wrote the original manuscript which was reviewed and modified by PA, FS and SEA. SVK, FS and SEA contributed to interpreting the results and the conceptualization of the study. FS performed the clinical evaluations of subjects and was responsible for the original cohort-data.

## Acknowledgements and funding

We thank Reeta Levo for her invaluable contribution as research nurse. We thank Ursula Lönnqvist, Pedro Bento Pereira, and Simo Soila for skillful technical assistance in the Helsinki laboratory. We thank Velma Aho for supplying and formatting the clinical metadata for the samples.

FS received funding from The Michael J. Fox Foundation for Parkinson’s Research, The Finnish Parkinson Foundation, the Academy of Finland (295724, 310835), Helsinki University Hospital (T1010NL101, TYH2018224, TYH2020335), and Hyvinkää Hospital (M6095PEV12).

## Competing interests

FS has patents issued (FI127671B & US10139408B2) and pending (US16/186,663 & EP3149205) that are assigned to NeuroBiome Ltd. FS is founder and CEO of NeuroInnovation Oy and NeuroBiome Ltd., is a member of the scientific advisory board and has received consulting fees and stock options from Axial Biotherapeutics. FS has received consulting and lecture fees from Orion, Abbvie, Herantis, GE Healthcare, Merck, and Teva. SEA has acquired a research grant from Weston Brain Institute. The funders have no role in the preparation of the manuscript.

## Data availability

Clinical data are not publicly available due to participant privacy and are available from the corresponding authors on reasonable request.

## Notes

### Clinical Trial

NCT01536769

### Author Declarations

The original gender, age and sex matched cohort was recruited for a pilot study in 2015 investigating PD and gut microbiota (Scheperjans et al 2015). The study was approved by the ethics committee of the Hospital District of Helsinki and Uusimaa. All participants gave written informed consent and the study was registered at clinicaltrials.gov (NCT01536769).

